# D-dimer dynamics in hospitalized COVID-19 patients: potential utility for diagnosis of pulmonary embolism

**DOI:** 10.1101/2020.09.21.20193953

**Authors:** Pau Cerdà, Jesus Ribas, Adriana Iriarte, José María Mora-Luján, Raquel Torres, Belén del Río, Héctor Ignacio Jofre, Yolanda Ruiz, Marta Huguet, Maria Paz Fuset, Sergio Martínez-Yélamos, Salud Santos, Núria Llecha, Xavier Corbella, Antoni Riera-Mestre

**Affiliations:** Department of Internal Medicine, Hospital Universitari de Bellvitge - Bellvitge Biomedical Research Institute (IDIBELL), L’Hospitalet de Llobregat, Barcelona, Spain; Department of Pneumology, Hospital Universitari de Bellvitge - Bellvitge Biomedical Research Institute (IDIBELL), L’Hospitalet de Llobregat, Barcelona, Spain; Department of Diagnostic Imaging, Hospital Universitari de Bellvitge - Bellvitge Biomedical Research Institute (IDIBELL), L’Hospitalet de Llobregat, Barcelona, Spain; Intensive Medicine, Hospital Universitari de Bellvitge - Bellvitge Biomedical Research Institute (IDIBELL), L’Hospitalet de Llobregat, Barcelona, Spain; Department of Neurology, Hospital Universitari de Bellvitge - Bellvitge Biomedical Research Institute (IDIBELL), L’Hospitalet de Llobregat, Barcelona, Spain; Department of Clinical Laboratory, Hospital Universitari de Bellvitge - Bellvitge Biomedical Research Institute (IDIBELL), L’Hospitalet de Llobregat, Barcelona, Spain; School of Medicine, Universitat Internacional de Catalunya, Barcelona, Spain; Faculty of Medicine and Health Sciences, Universitat de Barcelona, Barcelona, Spain

**Keywords:** pulmonary embolism, venous thromboembolism, D-dimer, anticoagulant therapy, thromboprophylaxis, Coronavirus disease 2019 (COVID- 19), Severe Acute Respiratory Syndrome Coronavirus-2 (SARS-CoV-2).

## Abstract

**Background:** A higher incidence of thrombotic events, mainly pulmonary embolism (PE), has been reported in hospitalized patients with COVID-19. The main objective was to assess clinical and weekly laboratory differences in hospitalized COVID-19 patients according to occurrence of PE.

**Methods:** This retrospective study included all consecutive patients hospitalized with COVID-19 who underwent a computed tomography (CT) angiography for PE clinical suspicion. Clinical data and median blood test results distributed into weekly periods from COVID-19 symptoms onset were compared between PE and non-PE patients.

**Results:** Ninety-two patients were included, 29 (32%) had PE. PE patients were younger (63.9 (SD13.7) vs 69.9 (SD12.5) years). Clinical symptoms and COVID-19 CT features were similar in both groups. PE was diagnosed after a mean of 20.0 (SD8.6) days from the onset of COVID-19 symptoms. Corticosteroid boluses were more frequently used in PE patients (62% vs. 43%). Median values [IQR] of D-dimer in PE vs non-PE patients were: week 2 (2010.7 [770.1-11208.9] vs 626.0 [374.0-2382.2]; *p*=0.04); 3 (3893.1 [1388.2-6694.0] vs 1184.4 [461.8-2447.8]; *p*=0.03); and 4 (2736.3 [1202.1-8514.1] vs 1129.1 [542.5-2834.6]; *p*=0.01). Median fold-increase of D-dimer between week 1 and 2 differed between groups (6.64 [3.02-23.05] vs 1.57 [0.64-2.71], *p*=0.003); ROC curve AUC was 0.879 (*p*=0.003) with a sensitivity and specificity for PE of 86% and 80%, respectively.

**Conclusions:** Among hospitalized COVID-19 patients, D-dimer levels are higher at weeks 2, 3 and 4 after COVID-19 symptom onset in patients who develop PE. This difference is more pronounced when the fold increase between weeks 1 and 2 is compared.

## INTRODUCTION

Coronavirus disease 2019 (COVID-19) is caused by severe acute respiratory syndrome coronavirus-2 (SARS-CoV-2). First reported in Hubei (China) at the end of 2019, COVID-19 rapidly spread worldwide and was declared a pandemic on March 11th, 2020 [1,2]. An unexpectedly high incidence of thrombotic events, mainly pulmonary embolism (PE), has been reported among patients hospitalized with COVID-19, particularly in intensive care units (ICU) [3-5]. Moreover, PE is a major worldwide health issue and the leading preventable cause of death in hospitalized patients [6].

Some studies have suggested that COVID-19 triggers a hypercoagulable state induced by hypoxia, immobilization, inflammation and cytokine storm syndrome, along with damage to endothelial cells [7]. More than one third of hospitalized patients with COVID-19 shows high D-dimer levels and D-dimer values are even higher in patients with severe COVID-19 than in those with mild disease [8-11]. Furthermore, endothelial dysfunction may cause blood coagulation and activation of platelet aggregation leading to vascular microthrombosis, which is considered crucial in the pathogenesis of the hypercoagulable state in COVID- 19 [12-14]. It cannot yet be ruled out that pulmonary endovascular filling defects found in these patients cause a local thrombotic phenomenon rather than true thromboembolism [12, 15].

The use of prophylactic doses of low-molecular-weight heparins (LMWH) is now widely recommended for patients hospitalized with COVID-19 [16, 17]. However, a high incidence of thrombotic events, mainly PE, has been reported despite the administration of standard thromboprophylactic doses [3, 5, 18]. In fact, some authors state that prophylactic-doses of LMWH might not be sufficient to deal with the COVID-19-related hypercoagulable state and that higher LMWH doses should be considered in hospitalized COVID-19 patients without documented venous thromboembolism [17, 19, 20].

There are two overlapping pathological subsets in COVID-19: the first is triggered by the virus itself and the second relates to the host response. Because of this dynamic clinical course, a staged clinical classification has been proposed [21]. This strategy is widely accepted, and clinicians generally assess COVID-19 in terms of weeks since onset of symptoms. However, the evidence on risk factors for PE in COVID-19 patients from this dynamic perspective is scarce and most studies are focused on ICU patients [3, 5, 22, 23]. The aim of our study was to compare clinical characteristics and laboratory data at different disease stages amongst patients hospitalized for COVID-19, according to the presence of PE detected on computed tomography (CT) scan.

## MATERIAL & METHODS

### Study design and patients

This was a retrospective, non-interventional study that included all consecutive patients admitted to the Hospital Universitari de Bellvitge (Barcelona, Spain) from March 1^st^ to April 24^th^, 2020 who met the inclusion criteria. During this period, a total of 2,558 patients attended the Emergency Department due to COVID-19 symptoms and 1,287 were admitted. The inclusion criteria were: 1) patients ≥18 years of age, 2) admission for COVID-19 pneumonia, and 3) chest CT angiography for clinical suspicion of PE during the study period. Given the 50%-80% sensitivity for SARS-CoV-2 real-time PCR, patients were also adjudicated as having COVID-19 if CT scan results were considered typical of the disease (i.e., extensive bilateral and peripheral ground glass opacities and/or alveolar consolidation), and if symptoms and/or blood test results were consistent with COVID-19 in the absence of an alternative diagnosis [24-26]. Patients with no contrast-enhanced chest CT scan were excluded, as were patients who were diagnosed with COVID-19 during a hospital stay for other medical conditions.

Data were obtained from routine daily practice and anonymized. Personal and clinical data collected for the study are in line with the Spanish Data Protection Act (Ley Orgánica 3/2018 de 5 de diciembre de Protección de Datos Personales). Informed consent was waived due to mandatory isolation measures in hospital care for these patients and because this was a retrospective study. The protocol was approved by the Ethics Committee of the Hospital Universitari de Bellvitge (Barcelona, Spain; approval number PR178/20). We followed the Strengthening the Reporting of Observational Studies in Epidemiology (STROBE) statement guidelines for observational cohort studies [27].

### Variables

The following parameters were collected: patient baseline clinical characteristics; comorbidities (such as chronic heart, lung or kidney disease); concomitant therapies; risk factors for venous thromboembolism (VTE); treatment received upon PE diagnosis; drug and dose of anticoagulant therapy; and outcomes during hospitalization. Disseminated intravascular coagulation (DIC) and sepsis-induced coagulopathy (SIC) were defined according to ISTH criteria [28].

Immobilized patients were those who had been immobilized for surgical or non-surgical reasons occurred within two-months prior to hospital admission. Active cancer was defined as a diagnosed malignancy, irrespective of administration of anti-cancer treatment. Chronic lung disease was defined as chronic obstructive pulmonary disease, asthma, interstitial lung disease or obstructive sleep apnea syndrome.

For normotensive patients with PE, stratification using the simplified version of the Pulmonary Embolism Severity Index (sPESI) was assessed [29]. All bleeding events were classified as ‘major’ according to ISTH criteria [30].

### CT protocol and Imaging Analysis

The routine protocol implemented in our department for patients with suspected PE is multidetector pulmonary CT angiography with 16-slice multi-detector CT (Toshiba Aquilion RXL) after intravenous injection of 60 ml iodinated contrast agent (Rovi Iomeron) at a flow rate of 4 ml/s, triggered on the main pulmonary artery. CT scan settings were 100 kVp, rotation time 5 s, average tube current 500 mA and pitch 1×16. All chest CT scans with patterns consistent with COVID-19 and presence of PE were reviewed by 2 expert thoracic radiologists blinded to patient status and clinical and laboratory test results.

### Blood tests

All consecutive blood tests were collected between admission and discharge. Routine hematological parameters were measured using Sysmex XN series instruments provided by Roche Diagnostics. These included hemoglobin levels and platelet, white blood cell, lymphocyte and neutrophils counts. Blood biochemistry parameters [urea, creatinine, glomerular filtration rate, aspartate aminotransferase, alanine aminotransferase, lactate dehydrogenase (LDH), creatine kinase (CK), troponin, albumin, procalcitonin, triglycerides, C-reactive protein, ferritin and interleukin-6] were measured using a Cobas c6000 analyzer and a Cobas c8000 analyzer (Roche Diagnostics, Mannheim, Germany). Coagulation parameters (prothrombin time, activated partial thromboplastin time and fibrinogen) and D-dimer levels were determined using an ACL TOP 750 System and ACL TOP 500 (Instrumentation Laboratory, Germany). For D-dimer, the upper normal limit was set at 250 μg/L, except for those patients aged over 50 years for whom we used the recommended age-adjusted cut-off (age × 10) [31].

All results were stored in the OMNIUM database, from SUNSET Technologies (Girona, Spain), and were recovered by the Cobas Infinity (Roche) software that integrates all laboratory information systems from our hospital. Since the primary objective of the study focused on the development of PE, blood tests were collected before PE diagnosis by CT, while those collected after diagnosis were disregarded. The widely accepted three-stage classification for COVID-19 was used, so blood test results were distributed into six periods of one week [21].

### Statistical analysis

All categorical variables are expressed as frequencies and proportions, and continuous variables as means with standard deviations (SD) or median and with interquartile range [IQR]. Normality of the distribution was assessed using the one-sample Kolmogorov-Smirnov test. For those variables that were not normally distributed, results are presented as medians with IQR. If more than one blood test of a given patient was available in the same weekly period, mean (SD) values were used. All variables were compared between patients with and without PE at thoracic CT angiography. We used Chi-square or Fisher’s exact tests to compare categorical data between groups. Two-tailed unpaired Student *t*-tests were used to compare normally distributed continuous data, and the Mann-Whitney ***U*** test for non-normally distributed continuous data comparisons. For those statistically different parameters, fold change from the upper normal limit at different weekly timepoints were also calculated comparing PE and non-PE patients. Furthermore, to assess dynamic differences between consecutive one-week periods, we also calculated fold increases by dividing the parameter values from a week by the parameter values from the previous week. The area under the curve (AUC) and the 95% confidence interval (CI) of the receiver operating characteristic (ROC) curve were obtained. The optimal cut-off points to predict PE were determined by Youden’s ***J*** statistic [32]. A two-sided p-value less than 0.05 was statistically significant. Analyses were performed using IBM SPSS Statistics, version 19.0 for the PC.

## RESULTS

### Baseline characteristics

Overall, 2,447 CT scans were performed during the study period and 101 patients who underwent contrast-enhanced chest CT for PE suspicion, were selected. Five patients were referred to another center and four were not hospitalized, so 92 patients were finally included in the study (**Fig. 1**). A positive RT-PCR result for SARS-CoV-2 was obtained from 91 patients; diagnosis was based on CT images, clinical symptoms, and blood test data consistent with COVID-19 in one case. Mean age was 68.1 (SD 13.2) years, most patients were male (74%) and Caucasian (90%). PE was objectively confirmed in 29 (32%) patients. Patients who developed PE were significantly younger (63.9 [SD 13.7] vs 69.9 [SD 12.5] years) and less frequently presented arterial hypertension (41% vs 64%). Clinical characteristics of patients according to PE diagnosis are shown in **Table 1**.

**Figure 1.**
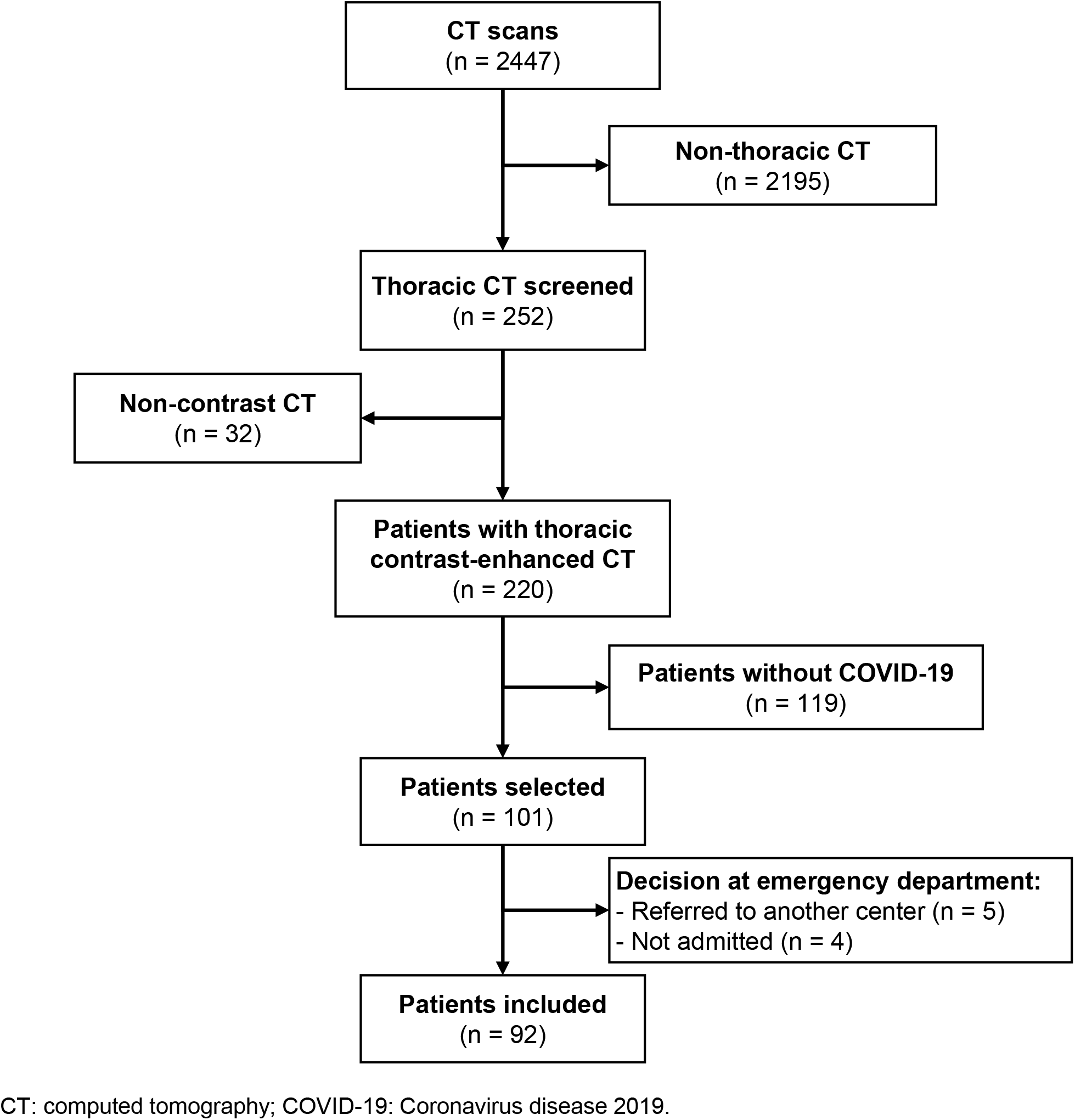
Patient selection.

**Table 1.** Clinical characteristics of patients according to the diagnosis of PE.

|  | **With PE**<br>**N=29** | **Without PE**<br>**N= 63** | **p-**<br>**value** |
| --- | --- | --- | --- |
| **Age, years; mean (SD)** | 63.9 (SD 13.7) | 69.9 (SD 12.5) | **0.039** |
| **Gender male, n (%)** | 23 (79%) | 45 (71%) | 0.424 |
| **Race Caucasian, n (%)** | 26 (90%) | 57 (91%) | 0.889 |
| **Body mass index; mean (SD)** | 30.2 (SD 5.3) | 28.7 (SD 4.2) | 0.199 |
| **Smoking behavior, n (%)** |  |  | 0.525 |
| Never smoker | 17 (59%) | 42 (67%) |  |
| Current smoker | 1 (3%) | 4 (6%) |  |
| Former smoker | 11 (38%) | 17 (27%) |  |
| **Alcohol history, n (%)** | 2 (7%) | 4 (6%) | 0.460 |
| **Comorbidity, n (%)** |  |  |  |
| Arterial hypertension | 12 (41%) | 40 (64%) | **0.047** |
| Diabetes mellitus | 5 (17%) | 22 (35%) | 0.084 |
| Dyslipidemia | 11 (38%) | 28 (44%) | 0.516 |
| Chronic lung disease | 6 (21%) | 14 (22%) | 0.868 |
| Chronic kidney disease | 2 (7%) | 11 (18%) | 0.215 |
| Atrial fibrillation | 0 | 2 (3%) | 0.490 |
| Ischemic heart disease | 1 (3%) | 5 (8%) | 0.661 |
| Cerebrovascular disease | 2 (7%) | 3 (5%) | 0.649 |
| Peripheral arteriopathy | 0 | 3 (5%) | 0.549 |
| **Pharmacological therapies at presentation, n (%)** |  |  |  |
| Antihypertensive medication | 12 (41%) | 37 (59%) | 0.121 |
| Antidiabetic medication | 4 (14%) | 19 (30%) | 0.085 |
| Statins | 8 (28%) | 24 (38%) | 0.325 |
| Antiplatelet therapy | 3 (10%) | 15 (24%) | 0.114 |
| Anticoagulant therapy | 1 (3%) | 4 (6%) | 1.000 |
| Hydroxychloroquine | 0 | 0 | - |
| **Risk Factors for Venous Thromboembolism (VTE), n (%)** |  |  |  |
| Previous thrombosis | 3 (10%) | 6 (9%) | 1.000 |
| Family history of VTE | 1 (3%) | 0 | 0.315 |
| Antiphospholipid syndrome | 1 (3%) | 0 | 0.315 |
| Immobility | 1 (3%) | 5 (8%) | 0.661 |
| Cancer | 1 (3%) | 5 (8%) | 0.661 |
| Use of contraceptives | 0 | 0 | - |

### In-hospital clinical characteristics according to PE

There were no statistically significant differences related to symptom type or time since onset before admission among PE and non-PE patients. On admission, both groups showed similar hemodynamics, oxygenation parameters and ventilation requirements. COVID-19-related CT findings were similar in both groups. All patients received similar experimental drugs for treating the disease, except for corticosteroid bolus, which was used more frequently in PE patients (62% vs 43%). None of the patients achieved ISTH criteria for DIC and less than 4% did so for SIC criteria (3% vs 3%). All patients received thromboprophylaxis from admission, except those who were already receiving anticoagulation therapy (3% PE vs 6% non-PE patients) and nine patients diagnosed with PE in the Emergency Department who immediately initiated anticoagulant treatment. Pharmacological thromboprophylaxis consisted of 3500 IU bemiparin OD or enoxaparin 40 mg OD or higher enoxaparin doses (including 40 mg BID or from 0.5-0.75 mg/kg BID). Interestingly, amongst the 15 patients who received higher enoxaparin doses for thromboprophylaxis, six patients developed PE while nine did not.

PE was diagnosed after 20 (SD 8) days from the onset of COVID-19 symptoms. Most PE were bilateral (52%) and the most proximal vessel involved was subsegmental in 10%, segmental in 34%, lobar in 31% and main pulmonary artery in 24%. PE features are shown in **Suppl Mat Table 1**. All patients received anticoagulant treatment with LMWH after PE diagnosis and no thrombolytic therapy or inferior vena cava filters were used. Major bleedings were significantly more frequent in PE patients (24% vs 8%); one non-PE patient with major bleeding was receiving higher doses of enoxaparin for thromboprophylaxis. Mortality was similar in both groups (27% vs 20%). COVID-19-related characteristics and in-hospital outcomes of patients according to PE are shown in **Table 2**.

**Table 2.** COVID19-related characteristics of the patients during hospital stay, according to the diagnosis of PE.

|  | **PE**<br>**N= 29** | **Non-PE**<br>**N= 63** | **p-value** |
| --- | --- | --- | --- |
| **Days of symptoms before admission, mean (SD)** | 9.6 (SD 5.2) | 7.5 (SD 5.8) | 0.104 |
| **Symptoms, n (%)** |  |  |  |
| Fever | 25 (86%) | 51 (81%) | 0.537 |
| Cough | 21 (72%) | 48 (76%) | 0.698 |
| Dyspneasubset | 18 (62%) | 42 (67%) | 0.667 |
| Arthromyalgia | 13 (45%) | 30 (48%) | 0.803 |
| Diarrhea | 11 (38%) | 22 (35%) | 0.780 |
| Anosmia | 1 (3%) | 6 (10%) | 0.426 |
| Dysgeusia | 2 (7%) | 7 (11%) | 0.714 |
| **Admission** |  |  |  |
| TAs | 136.7 (SD 21.4) | 128.3 (SD 18.7) | 0.061 |
| Sat O<sub>2</sub> | 93 (SD 5.7) | 94.1 (SD 4.9) | 0.337 |
| Sat O<sub>2</sub>/Fi O<sub>2</sub>, mean (SD) | 367.4 (SD 127.7) | 392.6 (SD 102.7) | 0.318 |
| **Length of stay (days), median (IQR)** | 20 (9.5 – 31.5) | 17 (11-30) | 0.791 |
| **COVID-related treatment, n (%)** |  |  |  |
| Hydroxychloroquine | 28 (97%) | 62 (98%) | 0.533 |
| Lopinavir/Ritonavir | 22 (76%) | 44 (70%) | 0.389 |
| Remdesivir | 1 (3%) | 3 (5%) | 1.000 |
| Tocilizumab | 13 (45%) | 24 (38%) | 0.541 |
| Corticosteroid bolus | 18 (62%) | 27 (43%) | **0.044** |
| Low-tapering corticosteroid therapy | 9 (31%) | 16 (25%) | 0.441 |
| **DIC score ≥ 5** | 0 | 0 | - |
| **SIC score ≥ 4** | 1 (3.4) | 2 (3.2) | 1.000 |
| **Thromboprophylaxis** | 28 (96%) | 59 (93%) | 1.000 |
| Higher doses enoxaparin | 6 (21%) | 9 (14%) | 0.484 |
| **Days of onset symptoms to CT** | 20 (SD 8) | 19 (SD 9) | 0.703 |
| **SatO<sub>2</sub>/FiO<sub>2</sub> at CT, mean (SD)** | 249 (SD 140) | 286 (SD 134) | 0.265 |
| **Pulmonary involvement in CT scan** |  |  |  |
| Bilateral pattern | 25 (86%) | 60 (95%) | 0.201 |
| Organizing pneumonia pattern | 11 (38%) | 20 (32%) | 0.560 |
| Ground glass pattern | 24 (83%) | 56 (89%) | 0.508 |
| Acute alveolar damage | 4 (14%) | 2 (3%) | 0.076 |
| Pneumo -thorax/-mediastinum | 2 (7%) | 1 (2%) | 0.233 |
| **Lower limbs deep venous thrombosis / Doppler US** | 6 (21%) / 6 | 6 (10%) / 12 | 0.054 |
| **Non-invasive / Invasive ventilation** | 11 (37%) | 11 (18%) | 0.087 |
| **Admission in Intensive care/step down unit** | 12 (41%) | 14 (22%) | 0.289 |
| **Bleeding** | 7 (24%) | 5 (8%) | **0.046** |
| Major | 5 (17%) | 4 (6%) | 1.000 |
| RBC transfusion | 4 (14%) | 4 (6%) | 0.576 |
| Embolization | 1 (3%) | 2 (3%) | 0.523 |
| **Mortality** | 8 (27%) | 13 (20%) | 0.485 |

*DIC, disseminated intravascular coagulation; SIC, sepsis-induced coagulopathy; CT, computed tomography; US, ultrasound; RBC, red blood cells*

#### Analysis of clinical laboratory data determinations associated with PE

Data from all blood tests were grouped by weeks from COVID-19 symptom onset and compared between PE and non-PE patients. Some differences were detected, but very few reached statistical significance (**Suppl Mat Table 2**). After discarding some clinically irrelevant variations, differences were only observed in median values [IQR] of D-dimer between PE and non-PE patients at week 2 (2010.7 [770.1 - 11208.9] vs 626.0 [374.0 - 2382.2]; *p* = 0.04); week 3 (3893.1 [1388.2 - 6694.0] vs 1184.4 [461.8 - 2447.8]; *p* = 0.03); and week 4 (2736.3 [1202.1 - 8514.1] vs 1129.1 [542.5, 2834.6]; *p* = 0.01) (**Fig. 2A)**. ROC analyses for D-dimer in these different weekly periods provided AUC values of 0.727 (*p* = 0.004), 0.743 (*p* = 0.003) and 0.746 (*p* = 0.01), respectively **(Fig. 2B)**. The optimal D-dimer cut-off points for PE according to Youden’s ***J*** statistic at weeks 2, 3 and 4 were 632 ug/L (*J*: 0.42), 2036 ug/L (*J*: 0.44) and 2271 ug/L (*J*: 0.42), respectively. Sensitivity and specificity results for these different cut-off points are shown in **Table 3**.

**Table 3.** AUC-ROC of D-dimer with various methods at different weeks from COVID19 symptoms onset.

|  | PE<br>Median (IQR) | No PE<br>Median (IQR) | AUC (CI) | p-value | Optimal Cut-off<br>(Youden's J) | Sensitivity | Specificity |
| --- | --- | --- | --- | --- | --- | --- | --- |
| **Blood levels (µg/L)** |  |  |  |  |  |  |  |
| Week 1 | 390 (200 – 1795) | 742 (340 – 2603) | 0.364 (0.140 – 0.587) | 0.223 | - | - | - |
| Week 2 | 2011 (770 – 11209) | 626 (374 – 2382) | 0.727 (0.605 – 0.849) | 0.004 | 632 pg/mL (J: 0.42) | 89% | 53% |
| Week 3 | 3893 (1388 – 6694) | 1184 (462 – 2448) | 0.743 (0.619 – 0.867) | 0.003 | 2036 pg/mL (J: 0.44) | 75% | 69% |
| Week 4 | 2736 (1202 – 8514) | 1129 (543 – 2835) | 0.746 (0.605 – 0.886) | 0.010 | 2271 pg/mL (J: 0.42) | 67% | 75% |
| Week 5 | 1916 (808 – 2672) | 981 (719 – 2071) | 0.600 (0.343 – 0.857) | 0.425 | - | - | - |
| Week 6 | 626 (341 – 2489) | 586 (368 – 1273) | 0.529 (0.206 – 0.852) | 0.845 | - | - | - |
| **Fold change from normal upper limit** |  |  |  |  |  |  |  |
| Week 1 | 1.56 (0.4 – 5.49) | 1.14 (0.72 – 4.43) | 0.48 (0.245 – 0.714) | 0.855 | - | - | - |
| Week 2 | 2.83 (1.37 – 26.16) | 0.92 (0.57 – 3.34) | 0.737 (0.608 – 0.866) | 0.004 | 1.41 (J: 0.39) | 77% | 63% |
| Week 3 | 6.93 (2.17 – 13.04) | 1.81 (0.69 – 3.5) | 0.735 (0.606 – 0.863) | 0.006 | 3.77 (J: 0.45) | 67% | 78% |
| Week 4 | 4.45 (1.88 – 12.16) | 1.66 (0.79 – 5.73) | 0.745 (0.594 – 0.897) | 0.013 | 1.83 (J: 0.37) | 82% | 55% |
| Week 5 | 3.36 (1.4 – 4.51) | 1.58 (1.13 – 4.12) | 0.611 (0.371 – 0.852) | 0.374 | - | - | - |
| Week 6 | 1.93 (0.67 – 3.74) | 1.2 (0.6 – 1.8) | 0.612 (0.277 – 0.947) | 0.457 | - | - | - |
| **Fold change from previous week** |  |  |  |  |  |  |  |
| Week 2 / Week 1 | 6.64 (3.02 – 23.05) | 1.57 (0.64 – 2.71) | 0.879 (0.748 – 1) | 0.003 | 2.87 (J: 0.66) | 86% | 80% |
| Week 3 / Week 2 | 3.25 (2.61 – 8.61) | 1.24 (0.71 – 4.06) | 0.668 (0.469 – 0.867) | 0.086 | - | - | - |
| Week 4 / Week 3 | 0.48 (0.26 – 3.14) | 0.91 (0.43 – 1.66) | 0.392 (0.147 – 0.637) | 0.317 | - | - | - |
| Week 5 / Week 4 | 0.80 (0.21 – 1.07) | 0.68 (0.49 – 1.73) | 0.453 (0.216 – 0.691) | 0.713 | - | - | - |
| Week 6 / Week 5 | 1.04 (0.24 – 1.67) | 0.71 (0.4 – 0.9) | 0.588 (0.200 – 0.977) | 0.557 | - | - | - |

Abbreviations: IQR: interquartile range; AUC: area under the curve; ROC: receiver operating characteristic curves; CI: confidence interval

**Figure 2.**
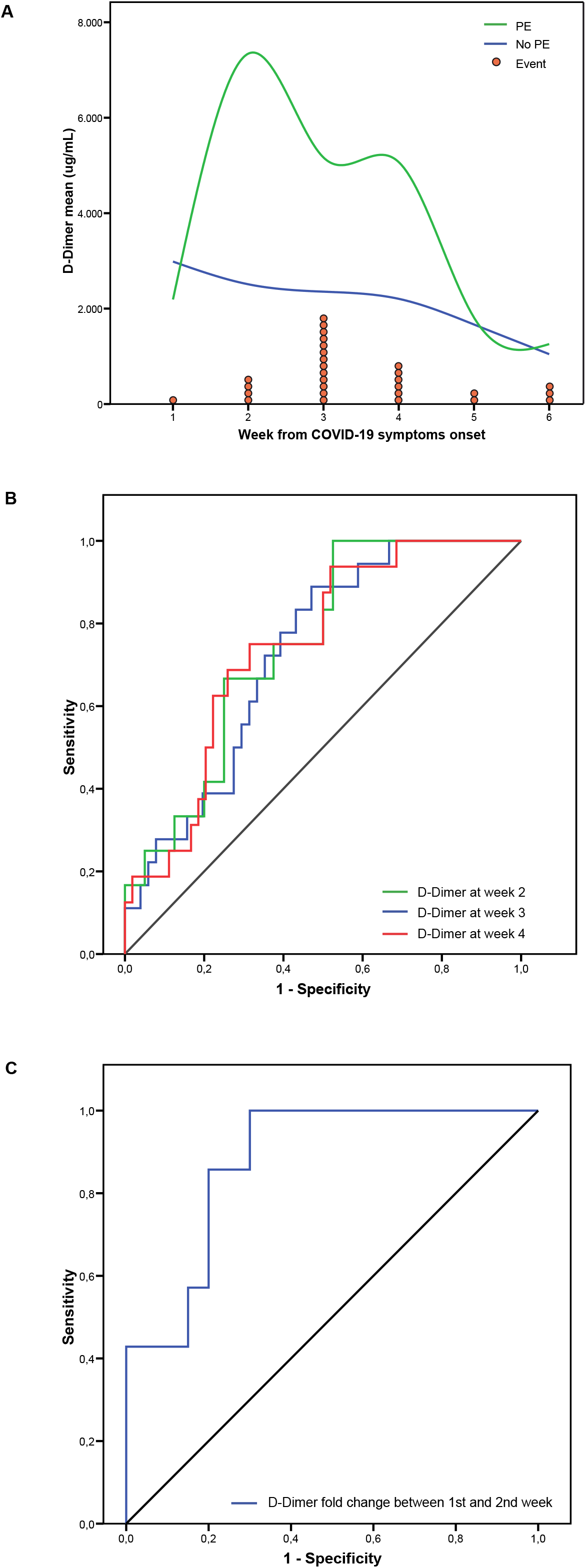
D-Dimer throughout time and its relationship with pulmonary embolism. **A**: D-Dimer blood level means through weeks from COVID19 symptoms onset, comparing patients with (green) and without PE (blue). Red circles represent each pulmonary embolism event in the week they were diagnosed. **B**: comparison of the different D-Dimer level ROC curves at week 2 (green), week 3 (blue) and week 4 (red). **C**: ROC curve of D-Dimer fold change between first and second week.

We also studied the magnitude of the difference in D-dimer levels compared to normal values. To this end, we calculated the D-dimer fold change from the age-adjusted upper normal limit at different weekly timepoints comparing PE and non-PE patients [28]. Median [IQR] fold changes were significantly higher in patients with PE than in those without PE, also at week 2 (2.83 [1.37 - 26.16] vs 0.92 [0.57 - 3.34], *p* = 0.004), week 3 (6.93 [2.17 - 13.04] vs 1.81 [0.69 - 3.5], *p* = 0.006) and week 4 (4.45 [1.88 - 12.16] vs 1.66 [0.79 - 5.73], *p* = 0.013). ROC analyses of these D-dimer fold changes at weeks 2, 3 and 4 yielded AUC values of 0.737 (p 0.004), 0.735 (*p* = 0.006) and 0.745 (*p* =0.013), respectively. The optimal D-dimer fold increase cut-off point for PE at weeks 2, 3 and 4 resulted in 1.41 (*J*: 0.39), 3.77 (*J*: 0.45) and 1.83 (*J*: 0.37), respectively. Sensitivity and specificity results for these cut-off points are shown in **Table 3**.

#### Analysis of clinical laboratory data fold increases between weeks associated with PE

As stated in methodology section, we calculated weekly fold increases for all blood test parameters and compared them between PE and non-PE patients. The only statistically significant and clinically relevant difference among PE and non-PE patients was D-dimer fold increase between the first and second week after COVID-19 symptoms onset (6.64 [3.02 - 23.05] vs 1.57 [0.64 - 2.71], *p* = 0.003). The AUC of the ROC curve was 0.879 (*p* = 0.003) and the optimal cut-off point according to Youden’s ***J*** statistic of D-dimer ratio between first and second week was 2.87 (*J*: 0.66). The prediction efficiency of all ROC analyses and sensitivity and specificity values are shown in **Fig. 2B and C** and **Table 3**.

## DISCUSSION

Though similar previous studies, this is the first one to report all blood tests performed in hospitalized COVID-19 patients by one-week periods, comparing groups with and without PE [22, 23, 33]. In a non-COVID-19 scenario, the negative predictive value of D-dimer testing is high, but the positive predictive value of elevated D-dimer levels is low. Therefore, in that setting, D-dimer testing is not useful for confirmation of PE, making pretest probability a key step in the diagnostic algorithm for PE [34]. However, symptoms of PE overlap with symptoms of COVID-19 pneumonia, so clinical suspicion of PE may be particularly challenging in patients hospitalized with COVID-19 illness. This is of clinical relevance, because COVID-19 patients often have severe hypoxemia, pulmonary hypertension due to hypoxic pulmonary vasoconstriction, or right ventricular failure, and an undiagnosed PE may be unrecoverable in such patients [4, 5, 18]. Although PE Wells score has been retrospectively used in a previous study, current clinical decision rules for PE diagnosis have not been validated for hospitalized COVID-19 patients [33]. Worryingly, most hospitalized COVID-19 patients show elevated D-dimer levels [22, 33]. Therefore, new D-dimer cut-offs that reflect the course of the disease over time and validation of previous predictive scores or design of specific scales are needed for assessing the clinical probability of PE in COVID-19 scenario.

We have determined the use of D-dimer levels in the COVID-19 scenario to calculate the clinical probability of PE, although this approach is the opposite to the conventional use of D-dimer to rule out this event [34]. Grouping these results into one-week periods helps clinicians identify the proposed disease stages for these patients and provides guidance to their findings [21]. We have found that after the viral response phase of the first week, D-dimer is the most useful factor for classifying hospitalized COVID-19 patients according to their risk of PE. Indeed, we have objectively confirmed the presence of higher D-dimer levels in weeks 2, 3 and 4 in patients with a diagnosis of PE during hospitalization. According to our findings, a 2.87-fold increase in D-dimer levels in the second week from COVID-19 symptom onset, compared to the first week, has a sensitivity and specificity for predicting PE of 86% and 80%, respectively.

Regarding clinical management in our series, the use of corticosteroid boluses was more common in patients presenting PE. Although a relationship between corticosteroid therapy and VTE has been previously described, early interventions aimed at reducing inflammation, such as dexamethasone, might help to prevent this hypercoagulable state [35]. In fact, treatment to counteract inflammation and cytokine storm syndrome has been related with lower mortality in COVID-19 patients [35, 36]. This is important because we have observed that PE was diagnosed mostly between 2-4 weeks after onset of symptoms, which means that PE occurs during or immediately after the systemic hyperinflammation stage, consistent with the ***thromboinflammation*** term [37, 38]. The inflammatory storm in COVID-19 can damage the microvasculature and cause endothelial dysfunction, which could trigger a hypercoagulable state [38, 39]. In fact, when endothelial dysfunction occurs, it leads to dysregulation of coagulation and complement and platelet activation, mainly mediated by α-thrombin [10, 38, 40]. According to our results, higher D-dimer values detected in PE patients compared to non-PE patients could reflect more intense endothelial damage and, consequently, a severe hypercoagulable state. Additional insight into possible individual factors related to PE developing during COVID-19 hospitalization is warranted.

SIC can be considered an earlier phase of DIC [28]. Although 3% of our PE patients met ISTH SIC criteria, none met DIC criteria. This suggests that severe COVID-19 is associated with coagulation derangements resulting in a hypercoagulable state rather than a consumption coagulopathy. Although specific drivers of this coagulopathy in COVID-19 are uncertain, it is known that SARS-CoV-2 can bind ACE2 and injure endothelial cells, leading to tissue factor expression and activation of the coagulation cascade. Elevated D-dimers may be a biomarker of this pathway [8, 41]. Indeed, we have found that those patients who developed PE during hospitalization showed higher D-dimer levels. These differences were statistically significant at weeks 2, 3 and 4, and also when compared with upper normal limit age-adjusted cut-off (age × 10) [31].

Once a thrombotic event such as PE occurs, anticoagulant treatment is recommended [17]. Controversy surrounds the doses of anticoagulants to be used in COVID-19 patients without a diagnosis of VTE. Although most authors recommend prophylactic anticoagulation, others suggest intermediate-dose parenteral medication or therapeutic anticoagulation [17]. Two retrospective studies suggested that anticoagulation treatment was associated with a reduced risk of mortality, but the efficacy and safety of higher doses of LMWH are still debated and may lead to undesirable events, such as those observed in our study. The use of intermediate or full therapeutic dose of anticoagulation agents needs to be addressed in randomized clinical trials, of which some are ongoing [19]. Moreover, because PE is usually diagnosed some days after admission, early thromboprophylaxis during hospitalization for COVID-19 patients is recommended and should be the first step to prevent VTE [17, 19]. However, since most of our PE patients were diagnosed between the week 2 and 4 after COVID-19 symptoms onset, it is important to have tools in clinical practice to identify patients at risk of developing PE despite prophylaxis. According to our results, weekly monitoring of D-dimer levels could help clinicians to select COVID-19 patients at higher risk for PE in whom a high clinical suspicion for PE should be addressed and higher anticoagulant doses might be evaluated, preferably in the context of clinical trials.

The present study has some limitations that should be mentioned. Firstly, the retrospective nature of the study, in which only patients with contrast-enhanced chest CT were considered, making the real PE incidence difficult to assess. Secondly, the relatively small size of the sample. However, we point out that this is the first study assessing all the commonly used blood test parameters in weekly intervals and that our results can be generalized to all COVID-19 patients, not only to ICU patients. Finally, external validation of our results is lacking. Despite that, our results could be helpful for the development of a new algorithm for PE diagnosis in hospitalized COVID-19 patients or to select those patients at higher risk of PE in randomized clinical trials aiming to assess the optimal dose of LMWH for thromboprophylaxis in this scenario.

In conclusion, D-dimer levels are higher at weeks 2, 3 and 4 after onset of symptoms in COVID-19 patients who develop PE during hospitalization, compared to those who do not develop PE. This difference is more pronounced when the fold increase between weeks 1 and 2 from symptom onset is compared. New weekly D-dimer cut-offs should be determined for assessing the clinical probability of developing PE in COVID-19 patients.

## Data Availability

The datasets used and/or analysed for the current study are available from the corresponding author on reasonable request.

## Acknowledgements

The authors thank the editorial support of Dr. Blanca Piedrafita at Medical Statistics Consulting for editorial support in completing this manuscript.

## Author contributions

PC and ARM conceptualized and designed the study; PC, AI, JR, JMML, RT, BR, HIJ, YR and ARM acquired the data; PC, AI, JR, JMML, RT, MH and ARM analyzed the data and interpreted the findings; PC, SMY and ARM performed statistical analysis; all authors contributed to the draft of the manuscript; JR, SMY, SS, NLL, XC and ARM performed critical revision of the manuscript; all authors approved the final version of the manuscript.

